# Complications Post Endovascular Abdominal Aortic Aneurysm Repair in Patients with Diabetes Mellitus: A Meta-analysis and Systematic Review

**DOI:** 10.1101/2023.03.25.23287746

**Authors:** EA Otify, M Mekki, J Borucki, K Dhatariya, P W Stather

## Abstract

**Background:** People with diabetes mellitus (DM) have higher long-term mortality following abdominal aortic aneurysm (AAA) repair than those without DM. However, whether this adverse outcome is directly related to their aneurysm is unclear.

**Aims:** To determine the rates of complications in people with and without DM post endovascular abdominal aortic aneurysm repair. Primary outcome data include AAA sac enlargement, reinterventions, endoleaks, post-operative AAA rupture and conversion to open surgical repair.

**Methods:** PubMed, Embase and Cochrane databases were searched for primary research studies between 2005 and 2023 according to PRISMA guidelines. Those undergoing AAA repair via endovascular aneurysm repair were included.

**Results:** Thirty-five studies were identified totalling 90,347 people in the control group, and 17,660 in the DM group. Those with DM had a lower rate of reintervention compared to controls (9.94% v 11.58%; OR 0.89, 95% CI [0.82-0.97]; P=0.005), however there was no significant difference in the rate of overall, type I or type II endoleaks (P=0.22, P=0.29, P=0.15 respectively).

People with DM were also less likely to have sac enlargement post AAA repair (9.66% v 11.27%; OR 0.79, 95% CI [0.68-0.93]; P=0.003). Additionally, people with DM had a significantly reduced rate of conversion to open surgery (2.11 % DM v 3.12% control; OR 0.80, CI [0.66-0.97]: P=0.02).

**Conclusion:** Reinterventions, sac enlargement post AAA repair, and conversion to open surgical repair were significantly lower in people with DM, however the cause for these differences remains unclear.

## Introduction

There is a well-established inverse association between diabetes mellitus (DM) and Abdominal Aortic aneurysm (AAA) growth ^[1]^, especially that of fasting glucose and aortic diameter, suggesting a relationship between glucose metabolism and AAA formation ^[2, 3]^. However, there is no association between serum glucose levels and aortic expansion ^[4]^.

Glycated haemoglobin (HbA_1c_) also has an inverse association with AAA growth rate in people with and without DM, suggesting a mechanistic relationship between long-lasting elevated blood glucose concentrations and AAA progression ^[4]^. These observations are contrary to the strong positive association of DM with other vascular diseases and their complications ^[5, 6, 7]^.

One of the first studies to highlight the diabetes paradox in AAA patients was an observational case-control study ^[8]^ utilising the AAA screening programme. This highlighted a significant reduction in AAA growth rates by 56% in patients using drugs used in the treatment of DM.

With the advancement of device technology and technique, more people are having Endovascular Aneurysm Repair (EVAR) compared to open surgical repair (OSR) ^[9]^. EVAR is associated lower early mortality; however, this approach has an increased long-term all-cause mortality, reintervention, and secondary rupture rates compared to OSR ^[10]^. Individuals with diabetes have higher rates of complications such as myocardial infarction, pneumonia, superficial surgical site infection, or pancreatitis following OSR ^[11, 12]^. Following EVAR, the range of complications differs in individuals, including endoleaks and endograft migration, and reintervention rates are typically between 16% to 30% ^[13]^. A previous meta-analysis has identified that diabetes is not a risk factor for type II endoleak ^[14]^ however there has been very little work done assessing the impact of DM on complications following EVAR. The aim of this systematic review and meta-analysis was to analyse the complications after EVAR in patients with DM.

## Methods

A final version of this protocol has been registered on PROSPERO in January 2023 prior to conducting the systematic review and meta-analysis (website: www.crd.york.ac.uk/prospero/ and ID: CRD42023379545). The review was conducted according to the Preferred Reporting Items for Systematic Review and Meta-analyses (PRISMA) guidelines ^[15]^.

### Eligibility criteria

Studies must have been written or translated into English, reporting on adults over 18 years old. Studies had to include people with diabetes who underwent EVAR, and have reported on at least one complication including reinterventions, sac enlargement, endoleak, post-operative AAA rupture or conversion to open surgical repair. Case reports, meeting abstracts, thoracic aortic aneurysms, laboratory-based studies, and reviews were excluded.

### Literature search

Two independent reviewers (E.O. and M.M.) performed a systematic search of the following 6 databases, PubMed, Medline, Embase, Cochrane Library, The Web of Science, and Google Scholar, for articles published between 2005 and 2023 using the search terms “Reintervention” AND “Abdominal Aortic Aneurysm” AND “Diabetes Mellitus”. “Endoleak” AND “Abdominal Aortic Aneurysm” AND “Diabetes Mellitus”. Researchers were contacted about published or unpublished trials identified through conferences or social media, to ensure all relevant studies were included. Ongoing trials were identified on www.clinicaltrials.gov. No contact was required to be made with authors. Zero publications required translation to be employed.

### Study identification and selection

A systematic search of the databases was undertaken by 2 reviewers (E.O. and M.M.) with duplicates removed, prior to screening the title and abstract of identified studies. Any discrepancies were discussed with the senior author. The remaining full-text articles were retrieved and independently reviewed for eligibility.

### Data extraction and quality assessment

Data extraction was undertaken by 2 reviewers using a standardised template. Any discrepancies were discussed. Descriptive, methodological and outcome data was input into a standard form. The reviewers extracted the following from each study: publication year, number of people with DM and controls, mean length of follow up, and outcomes reported from each of the studies to include endoleaks, reintervention, sac expansion, sac shrinkage, and conversion to open repair. The Newcastle-Ottawa scale was used to ensure the quality of non-randomised studies ^[16]^.

### Data synthesis and analysis

Meta-analysis was conducted of the adjusted risk estimates (where available) with use of the inverse variance method in Stata MP 17. Odds ratio was used to measure effect of each outcome. Statistical heterogeneity was assessed, with forest plots with an I^2^ ≤ 50% analysed using a Fixed Effects model (except AAA sac expansion where Random Effect model was used as each of the studies included had a different definition for AAA sac growth). For studies that published early and late results these were combined to determine an overall rate of each outcome. Sensitivity analyses were performed for all significant outcomes. Two sensitivity analyses were performed, one including all studies with >500 participants, and one including studies with a Newcastle Ottawa score of Good Quality.

## Results

The literature search identified 6165 articles, of which 3235 were excluded using automation tools. 308 articles were potentially suitable for inclusion in the review; 35 of these studies were eligible for inclusion (Figure 1). The characteristics of the included studies are outlined in Table 1.

**Figure 1.**
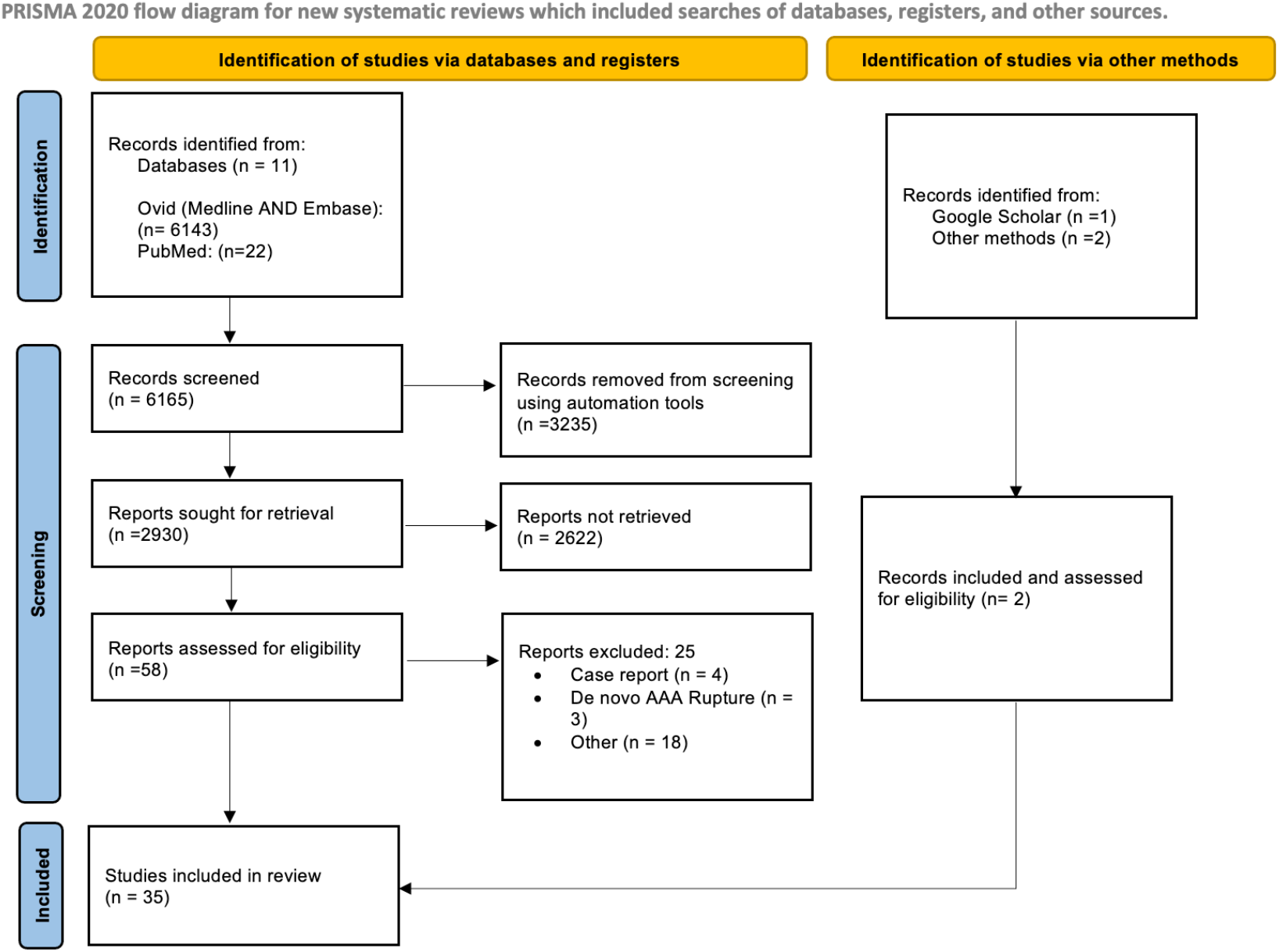
PRISMA Flow Diagram

**Table 1.** studies included in the meta-analysis

| Study | Study design, Country | No. of DM | No. of Control | Inclusion Criteria |
| --- | --- | --- | --- | --- |
| Png, 2017 | Retrospective Cohort, USA | 183 | 810 | Sac enlargement, Rupture, Reintervention and Type I, II, III and total Endoleak, conversion to open surgical repair |
| Taimour S., 2019 | Prospective Cohort, Sweden | 748 | 2630 | Reintervention |
| Leurs L.J., 2005 | Retrospective Cohort, Netherlands | 731 | 5286 | Reintervention, Type I, II, III Endoleak, conversion to open surgical repair |
| Hall M.R., 2017 | Retrospective Cohort, USA | 29 | 50 | Endoleak |
| Diehm, 2007 | Prospective Cohort, The Netherlands | 810 | 5573 | AAA sac enlargement, Reintervention, type I endoleak, conversion to open surgical repair |
| Takahara M., 2022 | Prospective Cohort, Japan | 226 | 703 | Aortic sac enlargement, Reintervention, total Endoleak |
| Praca, 2018 | Retrospective Cohort, Belgium | 57 | 191 | Aortic sac enlargement, Type II & total Endoleak |
| Seike Y., 2022 | Prospective Cohort, Japan | 2526 | 14573 | Type II Endoleak |
| Morisaki K., Matsubara Y., 2022 | Retrospective Cohort, Japan | 60 | 208 | Sac Shrinkage |
| Tseng HW,2022 | Retrospective Cohort, Taiwan | 36 | 155 | AAA sac enlargement and shrinkage after EVAR |
| Candell L, 2014 | Retrospective Cohort, USA | 441 | 1290 | Post-operative (EVAR) AAA Rupture |
| Hoshina K., 2019 | retrospective Survey, Japan | 4611 | 33397 | Sac enlargement and Reintervention |
| Lee JS, 2022 | Retrospective Cohort, South Korea | 60 | 294 | Aneurysm sac enlargement |
| Koole D., 2011 | Prospective Cohort, The Netherlands | 767 | 5570 | AAA sac enlargement and shrinkage after EVAR |
| Buijs RV, 2016 | Retrospective Case-control, The Netherlands | 10 | 50 | Type I Endoleak |
| Andersen R.M., 2018 | Retrospective Cohort, Denmark | 57 | 364 | endoleak after EVAR |
| Eden CL, 2020 | Retrospective Cohort, USA | 54 | 335 | Reintervention, Type II Endoleak |
| Morisaki K., Yamaoka T, 2017 | Retrospective Cohort, Japan | 29 | 182 | Sac enlargement and Type II Endoleak |
| Tan TW, 2016 | Retrospective Cohort, USA | 466 | 1936 | Type I Endoleak |
| Vaaramaki S, 2021 | Prospective Cohort, Finland | 30 | 189 | Sac Shrinkage, endoleak |
| Walker J., 2014 | Prospective Cohort, USA | 442 | 1294 | Type II Endoleak |
| Suarez G. LA, 2023 | Retrospective Cohort, Spain | 33 | 107 | Endoleak after EVAR |
| Wang Y., 2022 | Retrospective Cohort, China | 51 | 236 | Type II Endoleak |
| Major M., 2022 | Retrospective Cohort, USA | 54 | 335 | Type I Endoleak |
| Fujimura N, 2016 | Retrospective Cohort, Japan | 125 | 707 | Type II Endoleak |
| Shingaki M., 2018 | Retrospective Cohort, Japan | 25 | 148 | Sac Shrinkage |
| Kray J., 2015 | Retrospective Cohort, USA | 46 | 145 | Type II Endoleak |
| Suckow B.D., 2022 | Retrospective Cohort, USA | 4492 | 11445 | conversion to open surgical repair |
| Boniakowski A.E, 2016 | Retrospective Cohort, USA | 11 | 45 | Type II Endoleak |
| Sakaki M., 2020 | Retrospective Cohort, Japan | 96 | 454 | Type II Endoleak |
| Gibello L, 2020 | Prospective Cohort, Italy | 139 | 509 | Sac enlargement |
| Conrad M.F., 2009 | Retrospective Cohort, USA | 112 | 720 | Reintervention |
| Le T.B., 2018 | Retrospective Cohort, South Korea | 27 | 54 | Endoleak Post EVAR |
| Ichihashi S., 2019 | Retrospective Cohort, Japan | 71 | 362 | Type III Endoleak |
| Menges A.-L., 2022 | Retrospective Cohort, Switzerland | 5 | 30 | Reintervention |

### Reintervention

Meta-analysis of 9 studies including a total of 49,454 in the control group and 7,475 in the DM group identified significantly fewer reinterventions in those with DM (9.94% v 11.58%; OR 0.89, 95% CI [0.82-0.97]; P=0.005), (Figure 2).

**Figure 2.**
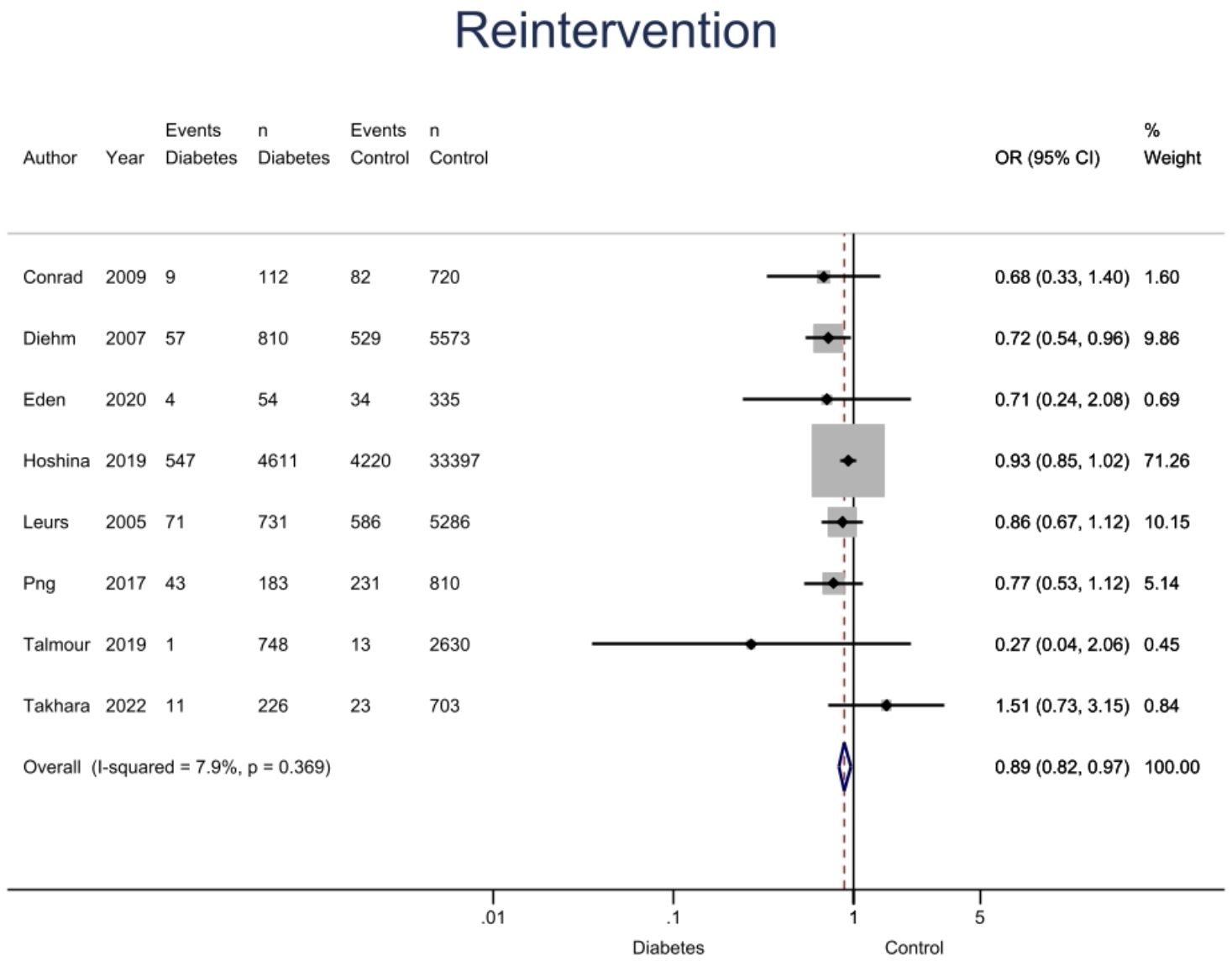
Forest Plot of reintervention rates post EVAR in those with DM and controls.

### Endoleaks

There was no significant difference in all endoleaks (28.33% DM v 31.97% controls; OR 0.91, 95% CI [0.53-1.58]; P=0.22). 6 studies reported on rates of type I endoleak and showed no significant difference (7.68% DM v 8.67% control; OR 0.91, 95% CI [0.77-1.08]; P=0.29) (Figure 3). 14 studies reported on rates of type II endoleak and showed no significant difference (26.41% DM v 26.76% control; OR 0.96, 95% CI [0.90-1.04]; P=0.33) (Figure 4). 3 studies reported on the rate of type III endoleak, identifying a lower rate of type III endoleak in those with DM (2.94% v 4.69%; OR 0.63, 95% CI [0.43-0.92]; P=0.02) (Figure 5).

**Figure 3.**
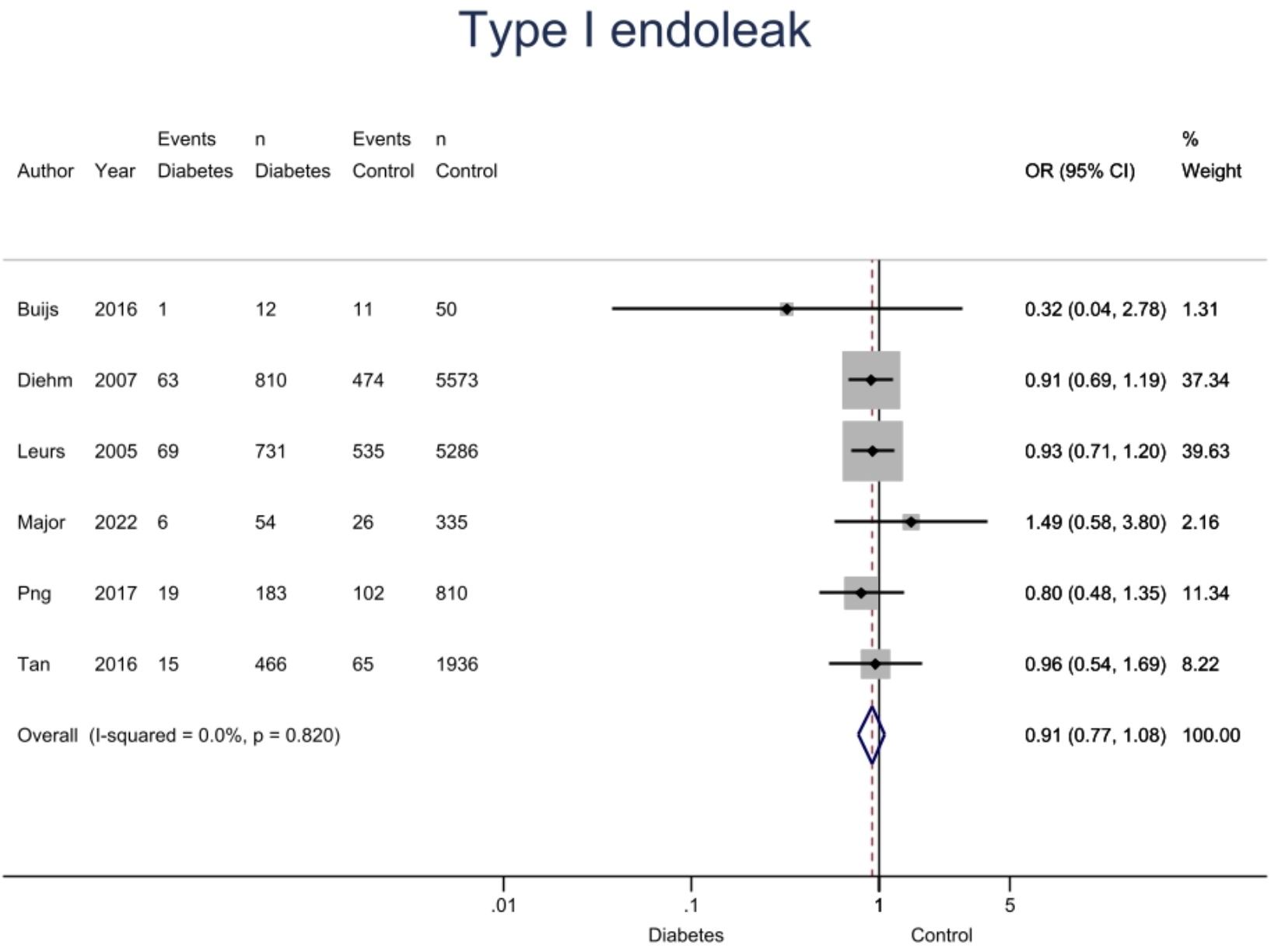
Forest Plot of Type I Endoleak post EVAR in those with DM and controls.

**Figure 4.**
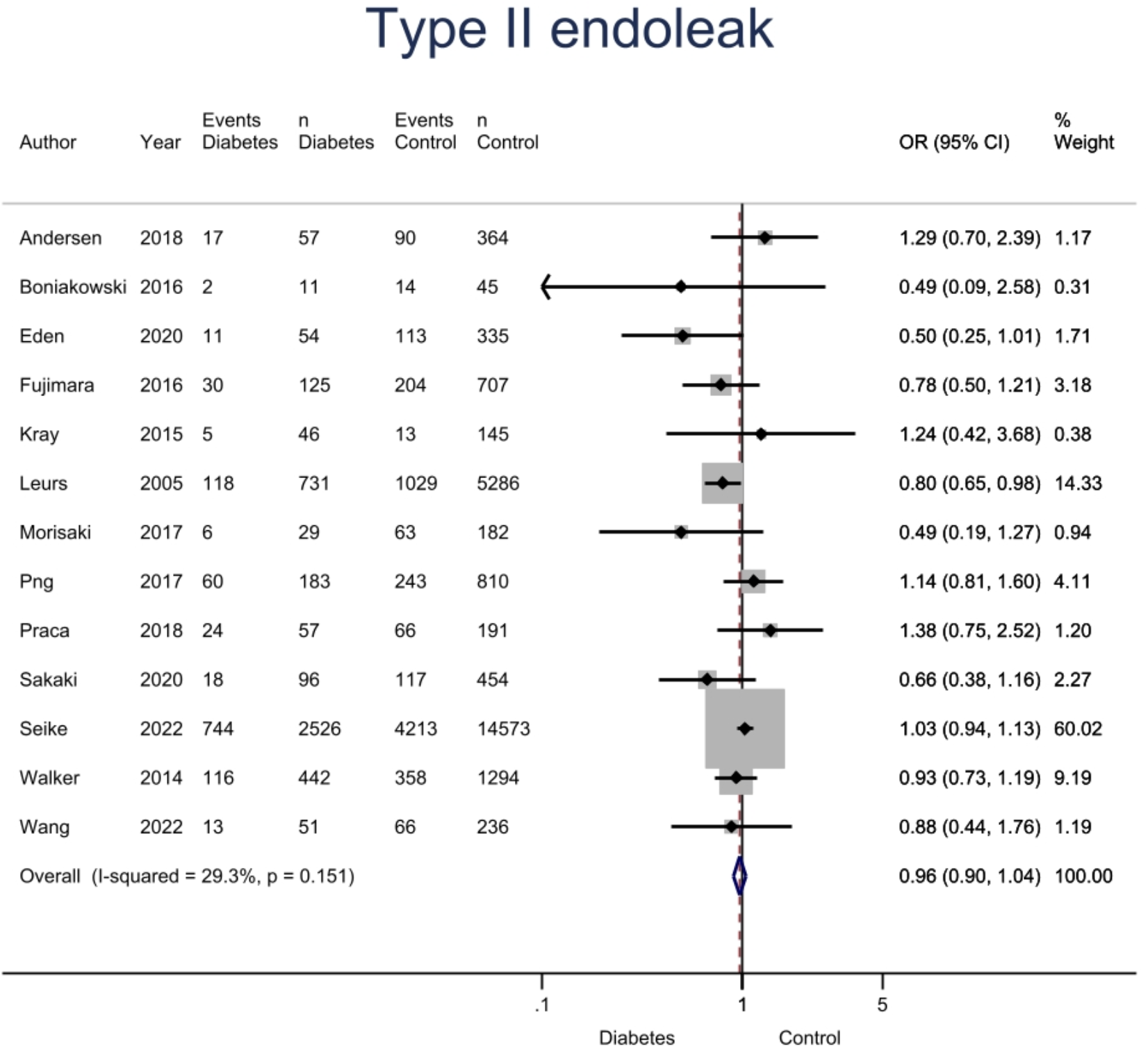
Forest Plot of Type II Endoleak post EVAR in those with DM and controls.

**Figure 5.**
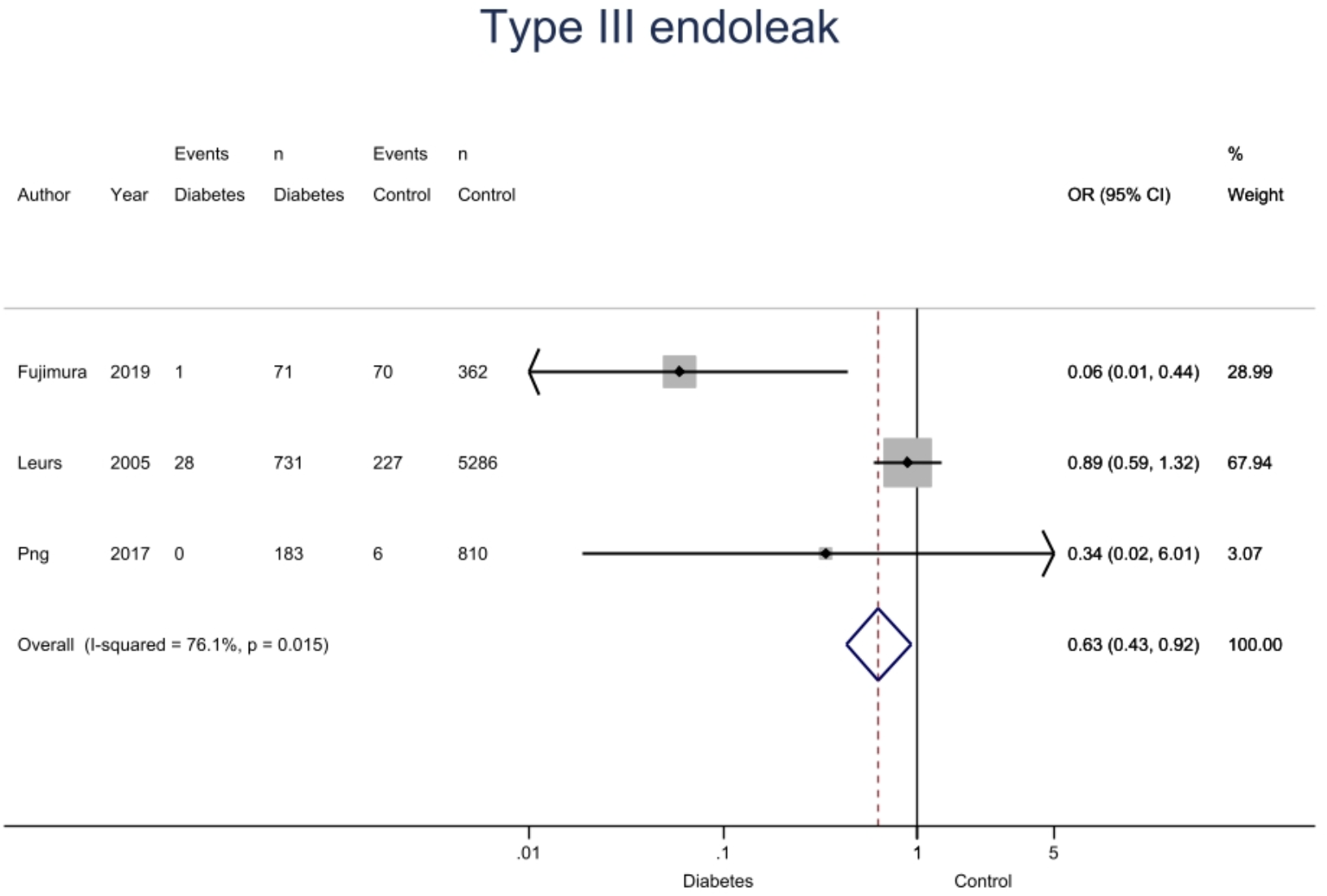
Forest Plot of Type III post EVAR in those with DM and controls.

### Sac change

Meta-analysis of 10 studies including a total of 47,384 in the control group and 6,918 in the DM group were assessed for AAA sac enlargement. Those with DM were less likely to have sac enlargement post AAA repair (9.66% DM v 11.27% controls; OR 0.79, 95% CI [0.68-0.93]; P=0.003) (Figure 6). There was no significant difference seen in the rate of sac shrinkage (19.80% DM v 17.81% controls; OR 0.97, 95% CI [0.89-1.09]; P=0.60) (Figure 7)

**Figure 6.**
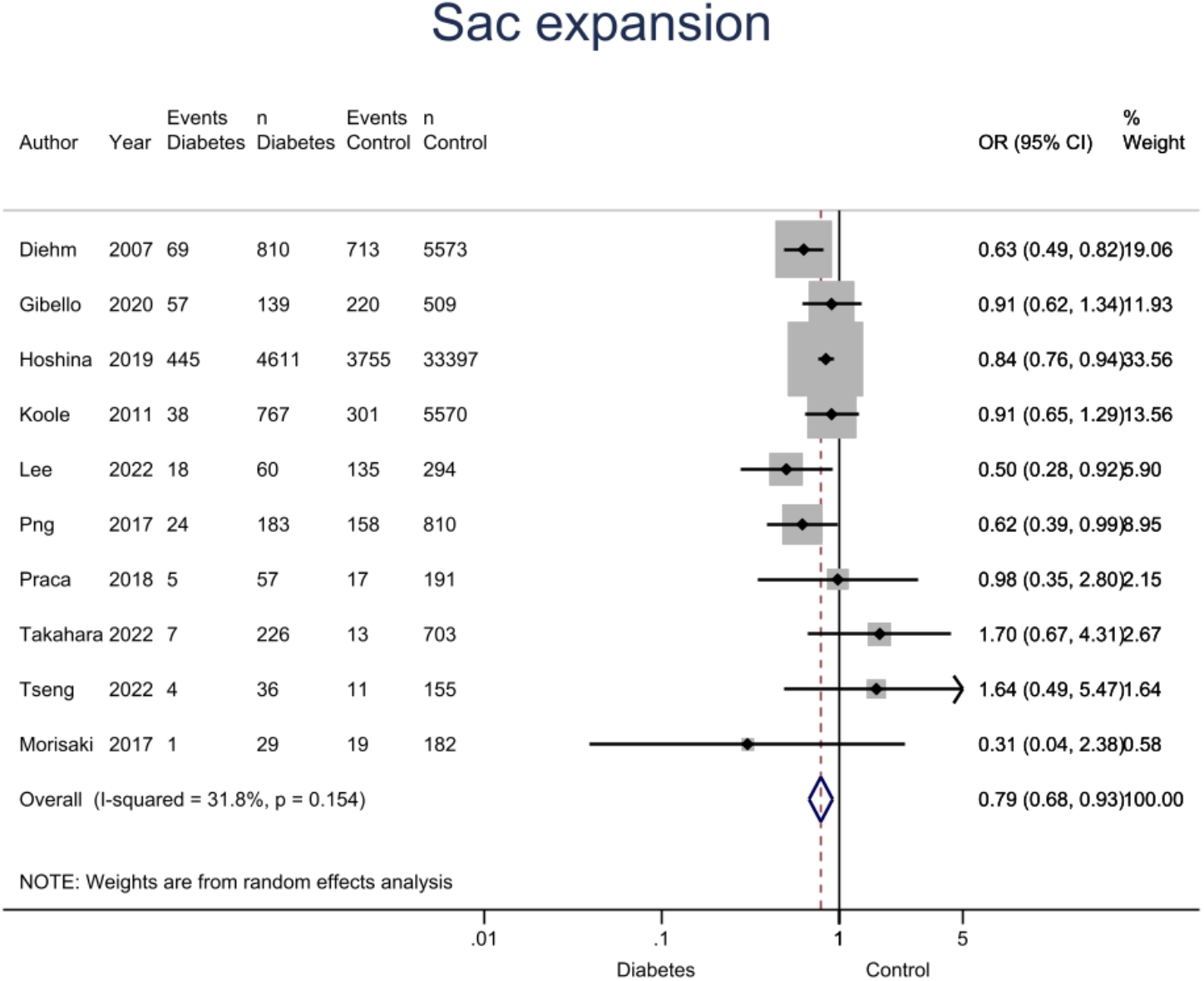
Forest plot of AAA Sac expansion in those with DM and controls

**Figure 7.**
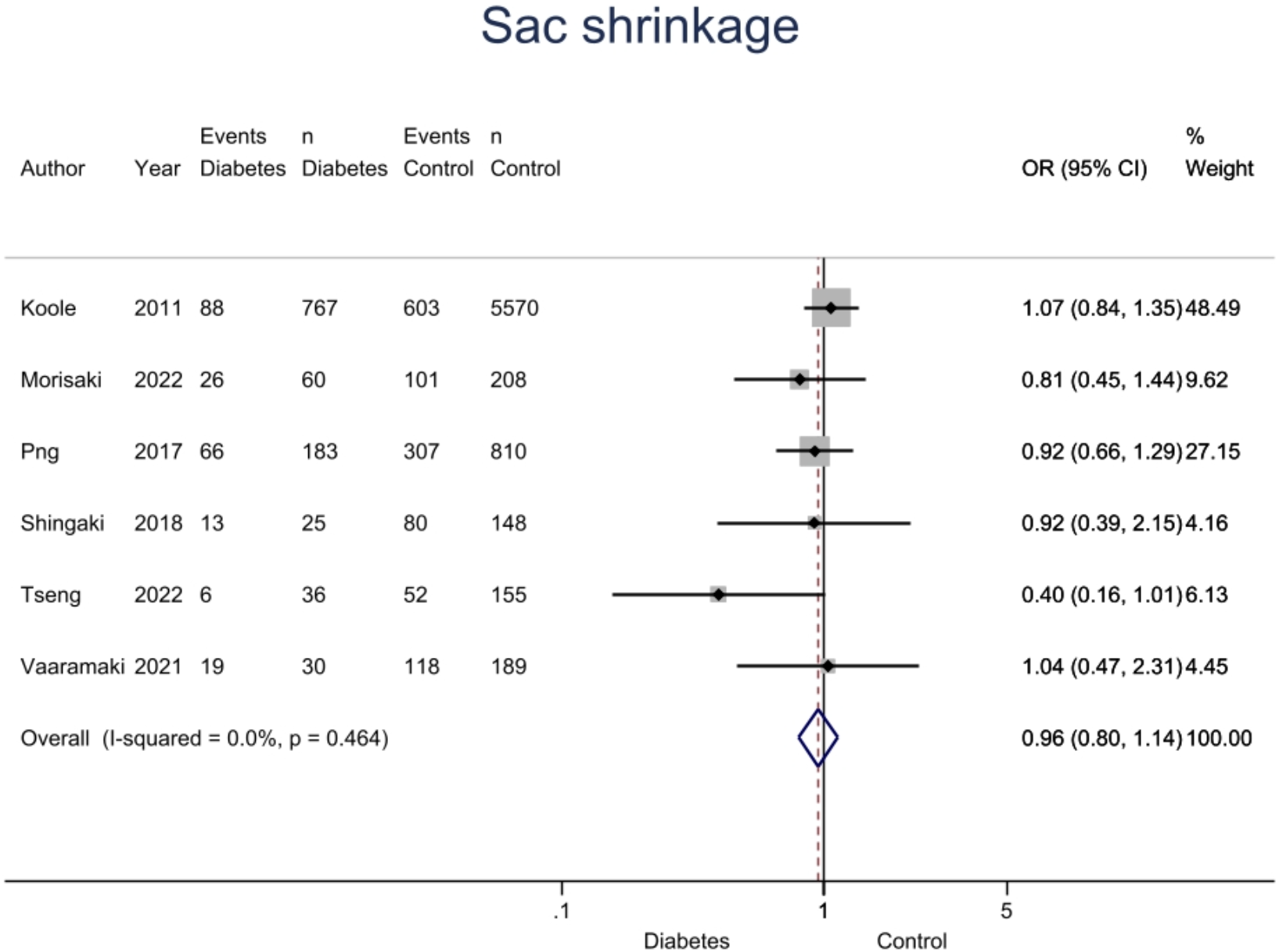
Forest Plot AAA Sac Shrinkage in those with DM and controls

### Rupture and conversion

There was no significant difference in the rate of AAA rupture post-EVAR (0.70% DM v 0.56% control; OR 1.18, 95% CI [0.87-1.61]; P=0.29) across 6 studies (Figure 8).

**Figure 8.**
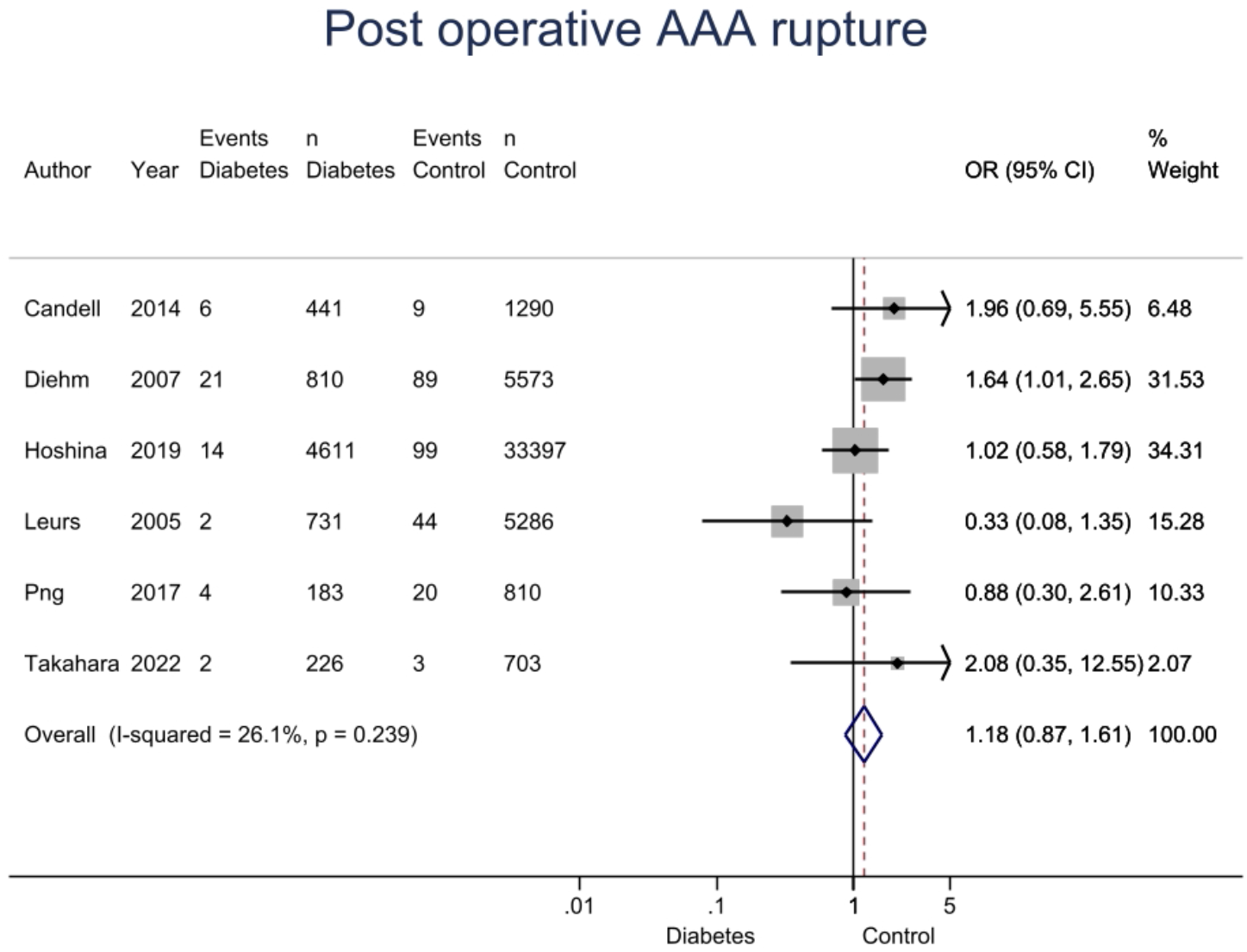
Forest plot AAA Rupture Post EVAR in those with DM and controls

Conversion from EVAR to open surgical repair was reported in 4 studies which was significantly less common in those with DM (2.11 % DM v 3.12% control; OR 0.80, CI [0.66-0.97]: P=0.02) (Figure 9).

**Figure 9.**
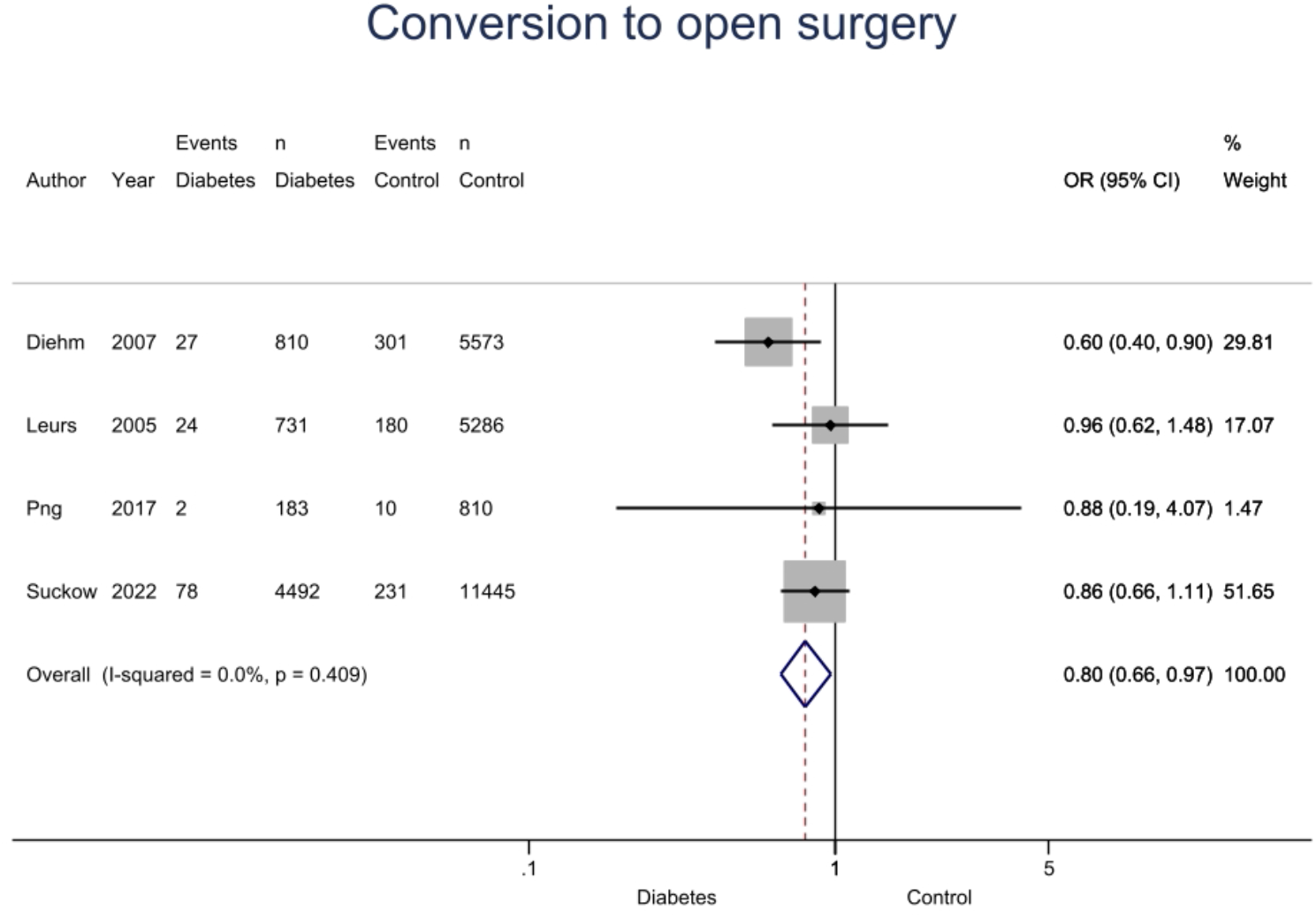
Forest plot Conversion to open surgical repair in those with DM and controls.

### Sensitivity analysis

Sensitivity analysis was undertaken for studies of >500 participants. This was possible for reintervention and AAA sac expansion. These results remained significantly different with lower rates in those with DM (P=0.006 and P=0.009 respectively) (Table 2).

**Table 2.** Sensitivity analysis of reinterventions and AAA sac expansion of sample size >500 participants

| **Outcome** | **No. Studies** | **DM (Event)** | **Total DM** | **Control (Event)** | **Total control** | **Total participants** | **OR, 95% CI** | **I<sup>2</sup>** | **P value** |
| --- | --- | --- | --- | --- | --- | --- | --- | --- | --- |
| Reinterventions | 7 | 739 | 7,421 | 5,684 | 49,119 | 56,540 | 0.89 [0.82-0.97] | 19% | 0.006 |
| AAA Sac Expansion | 6 | 640 | 6,736 | 5,160 | 46,562 | 53,298 | 0.80 [0.68-0.95] | 41% | 0.009 |

Sensitivity analysis was also undertaken utilising only studies that score as ‘good’ on the Newcastle-Ottawa Scale. The results for reintervention, sac expansion, and open surgical conversion remained significant (P=0.004, P=0.05 and P=0.04 respectively). There are results of type I or type II endoleak, AAA rupture post EVAR, and AAA sac shrinkage remained non-significant (Table 3).

**Table 3.** Newcastle-Ottawa Scale Quality Assessment Sensitivity analysis

| Outcome | No. Studies | DM (Event) | Total DM | Control (Event) | Total control | Total participants | OR, 95% CI | I<sup>2</sup> | P value |
| --- | --- | --- | --- | --- | --- | --- | --- | --- | --- |
| Reinterventions | 7 | 193 | 2,815 | 1,478 | 15,752 | 18,567 | 0.79 [0.67-0.93] | 0% | 0.004 |
| AAA Sac Expansion | 6 | 200 | 2,182 | 1,422 | 13,356 | 15,538 | 0.80 [0.64-1.00] | 35% | 0.05 |
| Type I Endoleak | 5 | 172 | 2,244 | 1,202 | 13,940 | 14,164 | 0.92 [0.78-1.09] | 0% | 0.33 |
| Type II Endoleak | 10 | 1129 | 4,229 | 6,296 | 23,651 | 27,880 | 0.98 [0.91-1.06] | 6% | 0.68 |
| Post-EVAR AAA rupture | 5 | 35 | 2,391 | 165 | 13,662 | 16,053 | 1.26 [0.87-1.83] | 34% | 0.22 |
| Conversion | 3 | 53 | 1,724 | 491 | 11,669 | 13,393 | 0.74 [0.55-0.99] | 19% | 0.04 |
| Sac Shrinkage | 4 | 186 | 1,005 | 1,108 | 6,717 | 7,722 | 1.01[0.84-1.22] | 0% | 0.89 |

### Publication bias

Funnel plots were reviewed for reintervention (Figure10), sac expansion (Figure11), and type II endoleak (Figure 12). As there were ≥ 10 studies included in each of those domains. Egger test was undertaken for reintervention, sac expansion and type II endoleak. These showed no evidence of publication bias, p values were 0.18, 0.77 and 0.27 respectively (Table 4).

**Figure 10.**
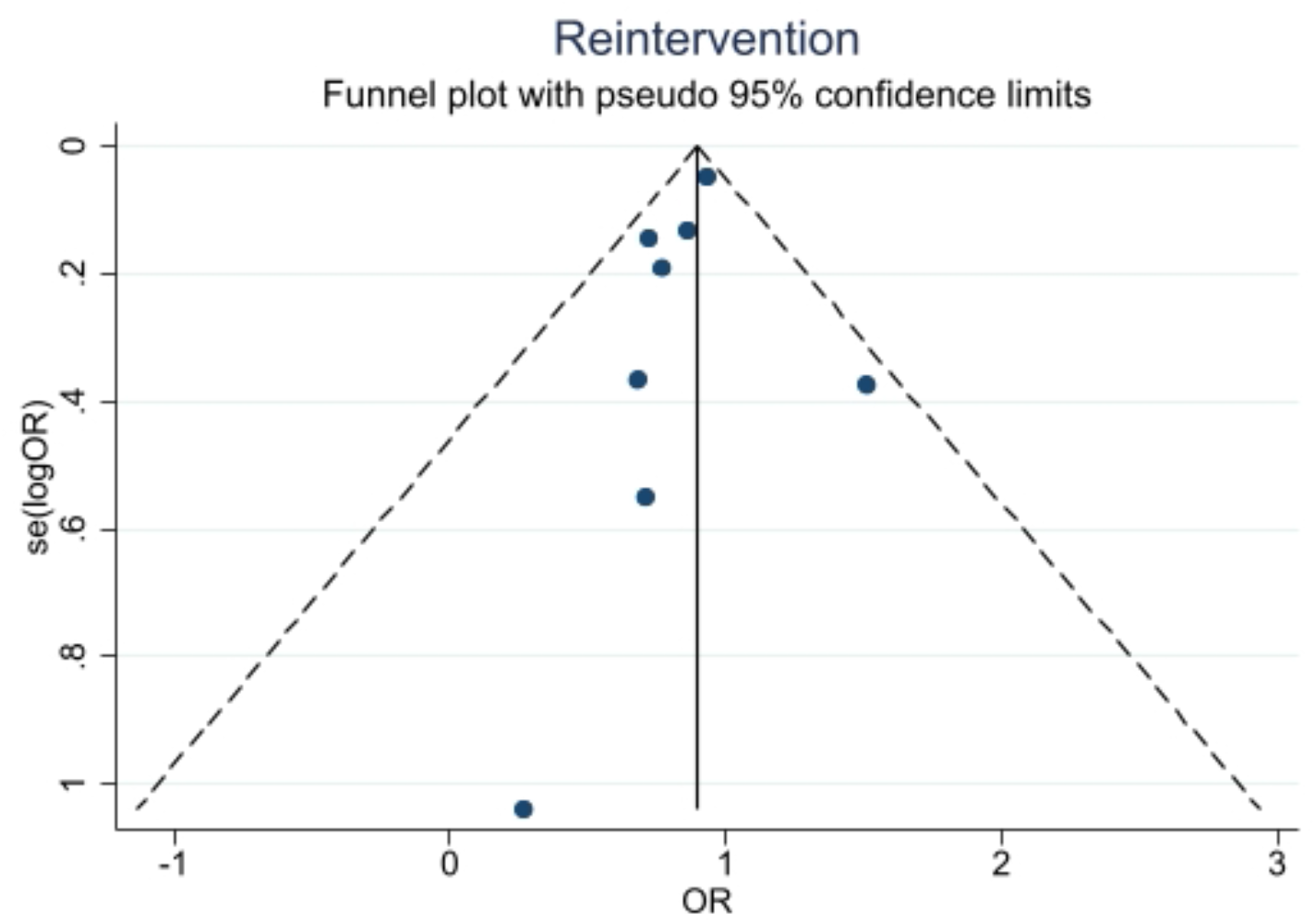
Funnel plot of AAA reintervention

**Figure 11.**
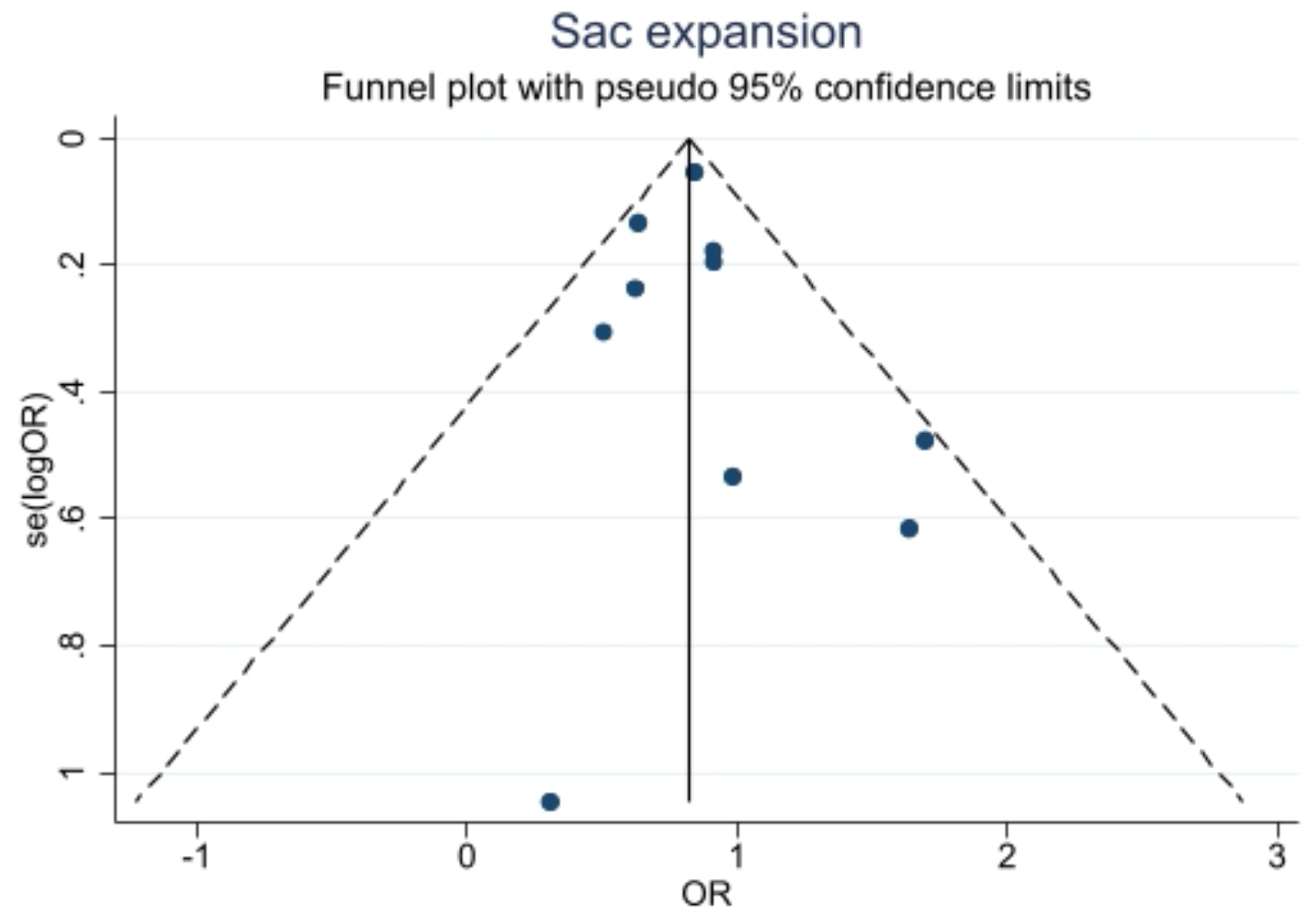
Funnel plot of AAA sac expansion

**Figure 12.**
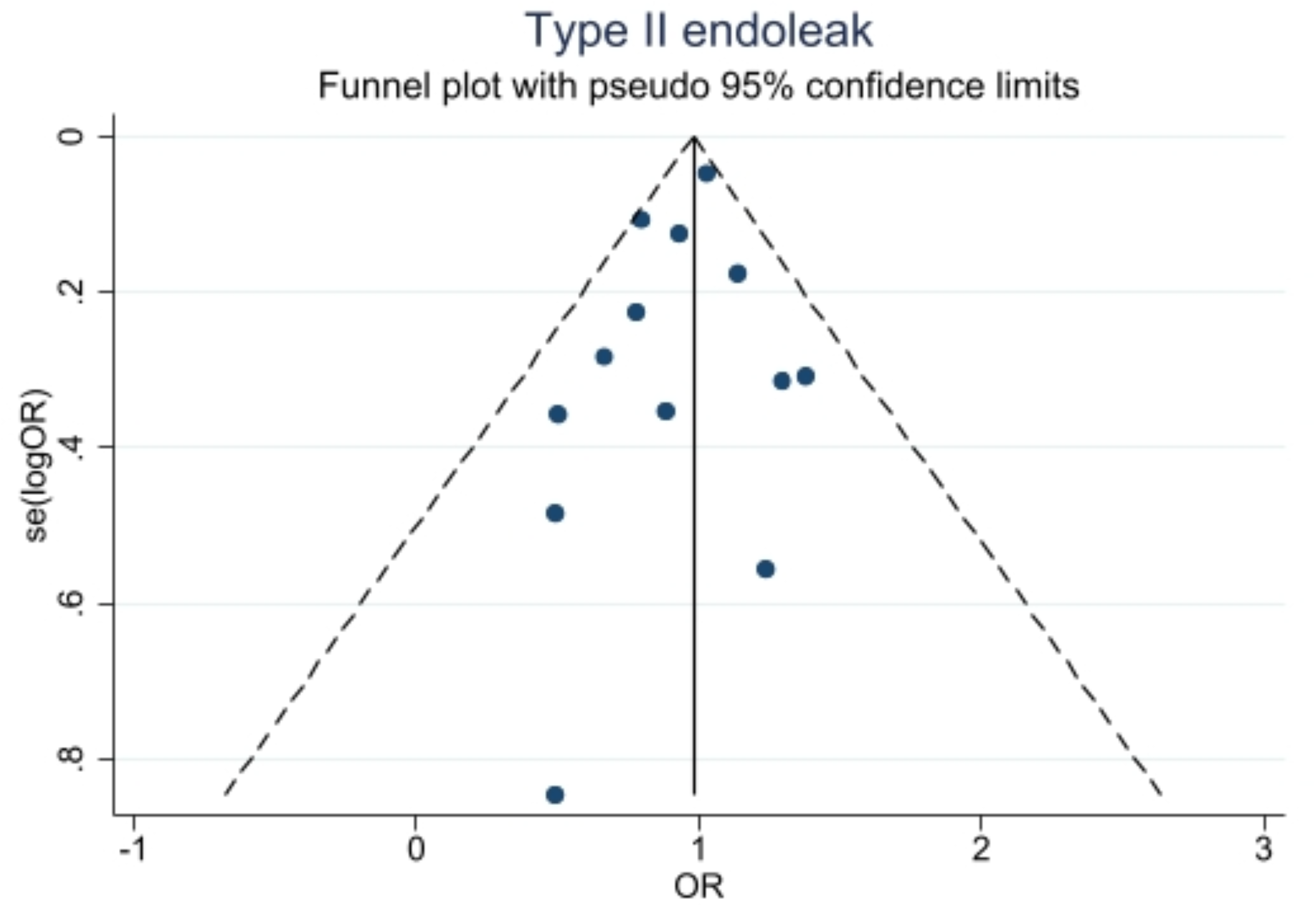
Funnel plot of type II endoleak

**Table 4.** Diabetes and AAA: Results from Eggers test

| Meta-analysis | Result | p |
| --- | --- | --- |
| Reintervention | no small-study effects | 0.1772 |
| Sac expansion | no small-study effects | 0.7694 |
| Type II endoleak | no small-study effects | 0.2698 |

## Discussion

This meta-analysis and systematic review has highlighted significantly lower rates of reintervention, sac enlargement, type III endoleak, and conversion to open surgical repair in people with DM undergoing EVAR. In addition, having diabetes was not associated with significant differences in overall rates of endoleak, type I or type II endoleak, sac shrinkage, or post-EVAR rupture compared to those without the condition.

People with diabetes are more likely to undergo EVAR as opposed to open surgical repair due to microvascular and macrovascular complications that are normally associated with diabetes making them more likely to be unfit for open surgical repair ^[17]^. Compared to those undergoing open AAA repair, those undergoing EVAR are more susceptible to late aneurysm-related complications due to progress of aortic degeneration and dilatation resulting in late aneurysm sac rupture, device failures, or endoleaks ^[18]^.

Reintervention impedes EVAR. This study identified 9 studies that reported reintervention rates in those with and without DM, of which only the study by Diehm et al showed a significant reduction in reinterventions ^[19]^. Meta-analysis however highlights this significant reduction which remains on both sensitivity analyses. Further analyses showed the rates of endoleaks, sac expansion, rupture and conversion were further subcategorised to try to determine the causal relationship between diabetes and reduced reintervention rates.

One of the main causes for reinterventions is endoleak. 6 studies reported on Type I endoleak none of which showed a significant difference similar to that of the meta-analysis. 14 studies reported on Type II endoleak of which only the study by Leurs et al ^[18]^ showed a significant reduction of type II endoleak in those with diabetes. This confirmed the findings by Guo et al ^[14]^ who undertook a meta-analysis of studies reporting on factors associated with type II endoleaks and found no association with DM. 3 studies reported type III endoleak, with the study by Fujimura et al ^[20]^ suggesting a significant reduction in type III endoleak in those with DM, which was confirmed on meta-analysis.

Previous work has highlighted a reduction in untreated AAA growth rates in people with DM ^[1]^. Sac expansion which could be equated to AAA growth was lower in those with DM post-EVAR in 4 out of the 10 studies included, with meta-analysis showing a significant reduction in sac expansion post EVAR in those with DM ^[17]^. These results remained significant on both sensitivity analyses. Leurs et al ^[18]^ reported that those with type 2 DM who were insulin-controlled had a significantly lower rate of endoleaks, which resulted in fewer secondary interventions compared to those taking oral hypoglycaemic agents or people without diabetes. One explanation for this could be because people with insulin-treated DM receive more intensive monitoring ^[21]^ and that insulin has an anti-inflammatory effect and reduces the pro-coagulant effect of hyperglycaemia ^[22]^. A case-control study suggested that DM causes microvascular disease and involves the vasa vasorum in the adventitia, which might not allow the leakage of pro-inflammatory cells into the arterial wall. Thus, the arterial wall not being able to expand ^[23]^. Unfortunately, it was not possible to do further subgroup analyses across the other studies depending on type of medication.

6 studies reported AAA rupture post EVAR, none of which showed a significant difference similar to that of the meta-analysis. 4 studies reported EVAR conversion to open surgical repair, of which only Diehm et al ^[19]^ showed a significant reduction in reinterventions in people with DM, however meta-analysis did identify a significant reduction in favour of those with diabetes.

Reintervention following EVAR can be a challenging clinical decision. Type I and Type III endoleaks have a significant risk of causing rupture, hence treatment is warranted. However, Type II endoleaks are often managed conservatively, with most clinicians not undertaking intervention unless the sac expands by at least 10mm ^[24, 25, 26]^. In addition, conversion to open surgical repair is a very complex procedure with a significant mortality ^[27]^. There may therefore be an underlying selection bias in some of these results as those with diabetes may be less likely to have their EVAR explanted due to their comorbidities. It is also unknown as to whether people with diabetes were more aggressively treated with EVAR, resulting in some grafts potentially being used outside of the manufacturers’ instructions for use, which would increase the risk of endoleaks, and post-EVAR rupture ^[28]^. Interestingly, although post-EVAR rupture was more common in those without DM, this was not significant, and the rate of Type 1 endoleaks was lower in those with DM, although this was also not significant. What remains unknown is the proportion of patients diagnosed with a type I or III endoleak who underwent intervention, and whether this varies between groups, which would affect post-EVAR rupture rates.

This study is limited by the variability in reporting. Whilst it is common to highlight the proportion of participants with diabetes, there was often no comparative analysis reported. In addition, not all outcomes were reported within each study. This is further highlighted by the lack of available data on iliac complications, with no reports on limb occlusion rates identified. Furthermore, most of the studies included did not differentiate between types of DM, or treatment. Given the distinct pathophysiological characteristics between type 1 and type 2 DM and the potential benefits of some glucose lowering agents, it is possible that relationships with AAA differ. The presence or absence of diabetes related co-morbidities were also not often reported, thus increasing the risk of selection bias, and potentially excluding those with more significant disease burden from inclusion into the studies.

The studies included were typically not able to eliminate the possibility of confounding variables in participants’ baseline characteristics because there were significant differences between those with or without diabetes in terms of comorbidities. Also, studies differ in the duration of the follow up which no doubt impact the outcomes. The Newcastle-Ottawa Scale Quality Assessment sensitivity analysis was used to try to explore this and account for such bias.

## Conclusion

This systematic review and meta-analysis of available data has shown that people with diabetes have lower AAA sac expansion, reintervention rates and conversion rates to open surgical repair post-EVAR. People with diabetes also have lower incidence of type III endoleaks compared to those without diabetes. Further work is necessary to establish why these differences occur.

## Data Availability

N/A

## Ethical approval

Not applicable.

## Informed consent

Not applicable.

